# Assessing the University Students’ Attitudes Toward Organ Donation in Jordan: A Cross-Sectional Study

**DOI:** 10.1101/2025.03.12.25323879

**Authors:** Redab Al-Ghawanmeh, Hindya O. Al-Maqableh, Mohammad Al-Qudah, Yasmeen Abu Sultan, Suhaib Alwreekat, Lujain Shaher AlShakhanbeh, Marylyn Ayoub, Malak M.M. Alhalawani, Dania Al-Omari, Zain Al-Tarawneh, Sara AL-Ajlouny, Aya Fargaly

**Affiliations:** Department of Pediatrics, Faculty of Medicine, The Hashemite University, Zarqa Jordan; Independent researcher, MCs (health services administration), Geo health Hub, JUST, Jordan; Department of Microbiology and Pathology and Forensic Medicine - Faculty of Medicine - The Hashemite University - Zarqa - Jordan; Jordan University of Science and Technology, Irbid, Jordan; MD. Medical Committee Member, Jordan Wrestling Federation, MOH); Medical Student, Hashemite University, Zarqa - Jordan; MD student, University of Balamand, Lebanon; Dania Al-Omari, BSc, Medical Education Coordinator, King Hussein Cancer Center, Jordan; BSc in Pharmacy, Amman Al-Ahliyah University, Amman, Jordan; Department of Health Services Management, Faculty of Economics, Yarmouk University, Irbid, Jordan

**Keywords:** Organ donation, university students, attitudes, willingness, Jordan

## Abstract

**Background:** Organ donation rates in Jordan are low despite improvements in healthcare. Cultural, religious, and familial beliefs influence attitudes toward donation. University students, as future societal leaders, are crucial in understanding these attitudes, yet research on this group in Jordan is limited. This study aims to assess university students’ attitudes toward organ donation in Jordan and identify factors influencing their willingness to donate.

**Methods:** A cross-sectional study was conducted with 1,548 university students from five universities in Jordan. Participants completed a self-administered questionnaire assessing demographics, attitudes toward organ donation (measured with a 5-point Likert scale), and willingness to donate. Statistical analyses included descriptive statistics, t-tests, ANOVA, and multiple linear regression.

**Results:** Seventy-two percent of students expressed a willingness to donate organs. The mean attitude score was 56.05/80, indicating moderate attitudes. Factors such as age, marital status, and willingness to donate significantly influenced attitudes. The strongest predictor of positive attitudes was willingness to donate.

**Conclusion:** University students in Jordan show moderate attitudes toward organ donation, with key influences from willingness to donate, age, and cultural beliefs. Targeted education and awareness campaigns are needed to address cultural barriers and improve organ donation rates.

## 1- Introduction

Organ transplantation is a life-saving intervention that greatly enhances the quality of life in patients with end-stage organ failure [1]. However, worldwide demand for organ donation continues to outpace supply, leading to a serious disparity in transplanting operations [2]. While some countries, such as Spain and the USA, have managed to develop effective campaigns for organ donation [3], others, notably Jordan, cannot balance supply and demand even with improvements in the infrastructure of healthcare [4]. Organ donation rates are generally low in many countries, especially in the Middle East, due in large part to a lack of adequate public awareness, cultural and religious attitudes, and legal limitations. These factors contribute to general confusion, particularly regarding permission from the family and post-mortem donation [5–6].

Organ transplantation has a relatively long history in Jordan, where, between 2011 and 2018, 1,553 transplant operations were carried out, comprising 1,643 kidney transplants and 110 liver transplants [4]. Though live donations are essentially confined to first-degree relatives, only a few of these involved deceased donors. Despite efforts to improve organ donation, the Jordan Center for Organ Transplantation Directorate of the Jordanian Ministry of Health registered only 30 new donors over six years [4], reflecting the ongoing challenge of increasing donation rates. The Middle East was one of the first to formally accept organ donation from both ethical and religious points of view. It established its formal recognition in the Council on Islamic Jurisprudence, held in 1987 in Amman, Jordan [7].

Families often oppose the donation decision, especially when it comes to brain death, which-while legally and medically recognized as death-is nonetheless difficult for many to conceptualize. This idea has also caused many controversies in countries where a brain-dead patient still shows physical appearance [8]. In the rest of the world, donation rates are strongly associated with public awareness and attitudes toward organ donation. In countries such as China, for instance, public knowledge about organ donation is rather high, but willingness to donate is rather low [9]. Similarly, in the Middle East, the knowledge of organ donation is at times more than the desire to help [4,10].

The attitudes of university students toward organ donation are important because they represent the future leaders of society. Research reports that while many individuals recognize the importance of organ donation, a significant number of people are hesitant due to concerns about body integrity, potential health risks, and religious beliefs [10,11,12]. Even with the continued growth in international studies related to organ donation, research concerning the attitudes of Jordanian university students. This disparity has inspired research into the opinions of Jordanian university students regarding organ donation and the factors that influence their decisions. The results from this study will be used to develop educational programs and legislation recommendations to improve organ donation in Jordan, while also addressing cultural, religious, and societal barriers.

Despite extensive research on organ donation attitudes worldwide, studies focusing on young adult populations, particularly university students in Jordan, remain limited. This gap is significant because university students represent a highly educated segment of society that can drive future changes in organ donation policies and social norms. Additionally, given their exposure to medical and ethical discourses, they can act as facilitators for awareness campaigns within their families and communities. Understanding their perspectives is essential for designing targeted interventions that address barriers unique to this demographic. This study, therefore, seeks to fill this gap by assessing not only their attitudes but also the predictors influencing their willingness to donate.

## 2- Method

### Study Design

This study employed a cross-sectional design to examine university student’s attitudes towards organ donation in Jordan. A self-administered questionnaire was distributed to students at five universities across Jordan, between 1st September 2024 to 30th December 2024.

### Study Population

The participants were university students from five universities in Jordan. Universities were selected based on geographic distribution and student diversity to ensure a representative sample of Jordanian university students. They were selected using random sampling techniques. surveys were randomly distributed to students across various faculties and academic years at the selected institutions. Inclusion criteria included students aged 18 years or older, currently enrolled in undergraduate or graduate programs at the selected universities.

### Sample size

The sample size was determined using an anticipated response rate, a confidence level of 95%, and an acceptable margin of error of 3%. Given a 20% non-response rate, the target sample size was set at 1,500 participants from the five universities. The sample comprised 1548 university students, exceeding the requisite representative sample size of 1,000 students for this population; consequently, a more representative sample will yield comprehensive insights into knowledge, attitudes, and willingness to donate among university students in Jordan.

### Questionnaire

This study used a self-designed questionnaire based on previous research [9], including three sections. The first component requested demographic information, including gender, age, educational background, and marital status. The second section included assessing individuals’ attitudes towards organ donation, with 20 questions on a 5-point Likert scale, from “strongly agree” to “strongly disagree.” Items 1 to 10 were evaluated using a scoring system where “strongly agree” received a score of 4 and “strongly disagree” received a score of 0. Conversely, for items 11 to 20, the score was inverted, assigning “strongly agree” a score of 0 and “strongly disagree” a score of 4. The attitudes were scored from 0 to 80, with higher scores indicating more favorable views towards organ donation. The third section included a single question about the willingness to donate organs. The answer was categorical (No, Yes).

### Pilot study

The questionnaire was evaluated for clarity and effectiveness using a pilot survey including a sample of 100 university students. The reliability and validity assessments of the instrument conducted using SPSS produced Cronbach’s alpha of 0.83, and pre-tested to ensure the validity. Feedback assisted in rephrasing, clarifying items, and enhancing item relevancy. Three experts in public health and psychology also reviewed the questionnaire for conceptual fit.

### Data collection

Data was gathered with a self-employed online questionnaire administered via Google Forms. The questionnaire was disseminated across multiple social media platforms to optimize reach and engagement from 1^st^ Nov 2023 to 31^st^ of May 2024. These platforms included social media networks like Facebook, WhatsApp, LinkedIn, Instagram, and student’s groups at the chosen universities. Furthermore, university emails were used to directly distribute the questionnaire link to students, and lecturers were approached to facilitate the dissemination of the survey among their pupils. As students themselves, the authors actively assisted in gathering responds by disseminating the survey link throughout their networks, ensuring adverse sample of participants.

### Ethical approval

the study was approved by the Institutional Review Board at Hashemite University, Zarqa, Jordan.

### Statistical Analysis

The categorical data were expressed in frequencies and percentages, the scale data expressed in mean and standard deviation. An independent t-test and one-way ANOVA were used to examine mean differences in attitudes toward organ donation based on students’ demographics. Multiple linear regression was utilized to predict factors associated with attitudes score. Alpha level set at 0.05 and any p-value less than alpha deemed statistically significant. SPSS Statistics for Windows, version 28.0 (SPSS Inc., Chicago, Ill., USA) was used to analyze data

## 3- Result

### Socio-demographic characteristics of study sample (N=1548)

A total of 1548 university students participated in the study. With a majority were female students (n=1179, 76.2%) compared to (n=369, 23.8%) were male students. Concerning marital status, the majority were single (n=1226, 79.2%) while a small proportion (2.0%) belonged to either divorced or widowed. Nevertheless, most students were between 19-25 years old (n=1064, 68.7%) compared to 484 students (31.3%) who were 26years or older. Additionally, 989 students (63.9%) were enrolled had medical background, while 36.1% were in non-medical majors. When a university students asked about their willingness to donate organs in the case of deceased donors, the majority (n=1123, 72.5%) expressed willingness, whereas 425 students (27.5%) were not willing to give their organs for other people. Table (1)

**Table (1).** Socio-demographic characteristics of study sample (N=1548)

| Variables | Category | Frequency | Percentage |
| --- | --- | --- | --- |
| Gender | Male | 369 | 23.8 |
|  | Female | 1179 | 76.2 |
| Marital status | Single | 1226 | 79.2 |
|  | Married | 291 | 18.8 |
|  | Others | 31 | 2.0 |
| Age | 19-25 years | 1064 | 68.7 |
|  | 26 and more | 484 | 31.3 |
| Education major | Medical majors | 989 | 63.9 |
|  | Non-medical majors | 559 | 36.1 |
| Willingness to donate your organs in the case of brain death | No | 425 | 27.5 |
|  | Yes | 1123 | 72.5 |

### The levels of university students’ attitudes toward organ donation

A total of 20 items distributed into three attitude domains were used to assess university students’ attitudes toward organ donation. The results in table (2) show that university students report a generally moderate attitude toward organ donation, with a total mean attitude score of 56.05 out of 80(*SD*=9.11). Moreover, the findings show that 45.5% of university students fall into the moderate attitude category, while 27.5% demonstrate low attitudes and 27.0% had high attitudes.

**Table (2).** association between students’ demographics, willingness to donate and attitudes toward organ donation.

| Variables | Category | N | Mean | SD | Test value | p-value |
| --- | --- | --- | --- | --- | --- | --- |
| Gender | Male | 369 | 55.70 | 9.04 | 0.855 | 0.393<sup>t</sup> |
|  | Female | 1179 | 56.16 | 9.13 |  |  |
| Marital status | Single | 1226 | 55.69 | 9.29 | 4.764 | **0.009<sup>F</sup>** |
|  | Married | 291 | 57.32 | 8.28 |  |  |
|  | Others | 31 | 58.32 | 8.19 |  |  |
| Age | 19-25 | 1064 | 55.37 | 9.44 | 4.406 | <0.001<sup>t</sup> |
|  | 26 and more | 484 | 57.55 | 8.15 |  |  |
| Education major | Medical majors | 989 | 56.08 | 9.41 | 0.198 | 0.843<sup>t</sup> |
|  | Non-medical majors | 559 | 55.99 | 8.56 |  |  |
| Willingness to donate your organs in the case of brain death | No | 425 | 50.19 | 9.25 | 16.969 | <0.001<sup>t</sup> |
|  | Yes | 1123 | 58.27 | 8.01 |  |  |

t: independent t test, F: one-way ANOVA

Among the attitude dimensions, the participants expressed the most positive views in the attitude of life view domain (*M*=3.18, *SD* =0.62), particularly recognizing organ donation as a life-saving and noble act (*M* =3.62, *SD* =0.67). Moreover, concerns had not persisted, for uncertainty about funeral arrangements and the perceived unnaturalness of donation. The attitude of family value dimension scored the lowest (*M* =2.47, *SD*= 0.74), showing a hesitation about family support and cultural perceptions. Moreover, the attitude of evaluation domain (*M*=2.58, *SD* =0.54) revealed skepticism regarding organ allocation, ethical concerns, and the potential risks of donation on recipient.

| No | Item description | Mean of 4 | SD |
| --- | --- | --- | --- |
|  | **Attitude of life view (items 1-8)** | **3.18** | **0.62** |
| 1 | Organ donation can save lives, showcasing an extraordinary act of kindness that benefits humanity. | 3.62 | 0.67 |
| 2 | If I die and donate my organs, a part of me lives on, establishing a new life with us as well. | 3.14 | 1.08 |
| 3 | I believe the body is a short-term vessel for the soul and mind and can be donated after death. | 3.18 | 1.04 |
| 4 | Organ donation complies with my beliefs. | 3.21 | 0.99 |
| 5 | I think signing an organ donation card is a noble and positive endeavor | 3.37 | 0.87 |
| 6* | If I donate my organs, I may not be able to have a funeral. | 2.95 | 0.89 |
| 7* | I prefer to die at home and not to donate my organs. | 2.94 | 0.90 |
| 8* | Donating organs is unnatural. | 3.07 | 0.85 |
|  | **Attitude of family value (items 9-11)** | **2.47** | **0.74** |
| 9* | My family would not support my organ donation after death. | 2.36 | 1.09 |
| 10* | Donating your family's organs may be viewed as disrespectful or selfish. | 2.32 | 1.09 |
| 11 | Donating organs according to elders' is an act of faithfulness | 2.72 | 1.06 |
|  | **Attitude of evaluation (items 12-20)** | **2.58** | **0.54** |
| 12* | I am not sure if the organ I donated will be transplanted to someone in need. | 2.36 | 1.12 |
| 13 | Organ donation can save lives, but NOT always. | 2.90 | 0.81 |
| 14* | I feel that my age or health will not allow me to donate an organ after death | 2.62 | 0.96 |
| 15* | If I donate organs, my body may not be respected. | 2.58 | 1.13 |
| 16* | If I consent, the physicians may remove my organs before verifying my death. | 2.60 | 1.12 |
| 17* | Organ donation could lead to developing certain diseases in a recipient | 2.49 | 1.03 |
| 18 | Donating an organ would lead to becoming close with the recipient and stay in touch with them | 2.75 | 1.06 |
| 19 | Maintaining a connection with the recipient of the donated organ creates feelings of kinship and connection | 2.70 | 1.06 |
| 20 | Families of organ donors can receive appropriate assistance during periods of financial difficulty. | 2.18 | 1.08 |
|  | **Total attitude score toward human organ donation (mean score out of 80)** | **56.05** | **9.11** |
|  | **Levels of attitude based on interquartile** | **N** | **%** |
|  | Low attitude (0-50) | 425 | 27.5 |
|  | Moderate attitude (51-61) | 705 | 45.5 |
|  | High attitude (62-80) | 418 | 27.0 |

\* Reversed items: 0=strongly agree, 1=agree, 2=neutral, 3=disagree, 4= strongly disagree

### Association between students’ demographics, willingness to donate and attitudes toward organ donation

Table (2) shows that marital status had a significant impact on students’ attitudes score *F* (2,1545) =4.761, *P*=0.009. The Games-Howell post hoc test demonstrates that only married students (*M*=57.32, *SD*=8.28) reported significantly higher attitudes scores compared to single students (*M*=55.69, *SD*=9.29, *P*=0.010). Moreover, individuals with age group 26years and above (*M*=57.55, *SD*=8.15) reported significantly higher attitudes compared to those between 19-25years old (*M*=55.37, *SD*=9.44),*t (1546) =4.406, P*<0.001). Additionally, students who expressed willingness to donate (*M*=58.27, *SD* =8.01) exhibit significantly higher attitude scores compared to those who were unwilling to donate (*M*= 50.19, *SD* = 9.25), *t* (*1546) =16.969, P<0.001).* The students’ gender and university specialty had no significant impact on attitudes score toward organ donation *P>0.05*

A prediction model was constructed based on variables that showed a significant association with attitudes toward organ donation namely (marital status, age and willingness to donate). Utilizing backward multiple linear regression analysis, only age and willingness to donate were retained in the final model *F* (2,1545)=147.201, *P*<0.001and both explained 16.0% of total variance in attitudes toward organ donation.

The strongest predictor was willingness to donate, as students who expressed willingness to donate reported an average attitudes score 7.921 units higher compared to those who were unwilling to donate (*B*=7.921, *t*=16.480, *P*<0.001). Moreover, students aged 26 years and older reported higher attitudes toward organ donation by 1.095 units compared to those aged 19-25years old (*B*=1.095, *t*=2.366, *P*=0.018). Table (3)

**Table (3).** predictors for university students’ attitudes toward organ donation.

| Predictors | Unstandardized Coefficients | Standardized Coefficients | t-value | p-value | 95.0% Confidence Interval for B |
| --- | --- | --- | --- | --- | --- |

|  | B | Std.<br>Error | Beta |  |  | Lower<br>Bound | Upper<br>Bound |
| --- | --- | --- | --- | --- | --- | --- | --- |
| Willingness to donate your organs in the case of brain death (Yes vs no) | 7.921 | 0.481 | 0.388 | 16.480 | 0.001 | 6.978 | 8.864 |
| Age group (26 years and more vs 19-25years) | 1.095 | 0.463 | 0.056 | 2.366 | 0.018 | 0.187 | 2.002 |

DV: attitudes toward organ donation, $F(2,1545)=147.201, P<0.001$ , R-square 0.160, adjusted R-squared=0.159

## 4- Discussion

With most respondents as medical students, 72.5% of them showed positive attitudes about organ donation. This can be attributed to their medical background, which facilitates a better understanding of medical ethics and transplantation [13]. Meanwhile, 27.5 % of students showed refused to participate which is an indication of the fact that issues related to bodily integrity and post-mortem donation are more influenced by the cultural and religious beliefs. These factors can therefore provide insight into the generally moderate approach that university students take toward organ donation [14]. Other factors deterring potential donors include misinformation about the distribution of organs, ethical concerns, and fears about the removal of organs too early [15]. Limited personal experience with organ donation, particularly among younger students, relates to lower degrees of support. Without direct access to transplanting events, students may be less likely to view organ donation in a favorable light [16].

Moreover, the findings showed that university students possess a moderate attitude towards organ donation, with a mean score of 56.05 out of 80, with a SD = 9.11. Nearly half of the students exhibited a moderate attitude (45.5%), whereas 27.5% displayed low attitudes and 27.0% high attitudes. The findings indicate a nuanced perspective on organ donation, with a considerable number of students holding reservations even while acknowledging its potential to save lives [17]. Several studies in the UK [18] Syria [19] and Jordan [10,20,21,]similarly identified cultural, ethical, and personal factors influencing attitudes toward organ donation, which also in line with the findings of our study.

The strong positive responses in the attitude to life view area indicate a belief that significant numbers of students value the humanitarian and saving-lives aspects attributed to organ donation. However, the residual apprehensions concerning funerals and feelings of unnaturalness of donations imply deep-seated social fears. Ambiguity about body integrity after death is likely to muddy these confusions [22]. All this reluctance in having frank discussions about death further limits proactive organ donation, hence perpetuating existing misconceptions. Misconceptions about brain death, in addition to burial customs and bodily integrity, are greatly shaped by cultural and religious views toward organ donation in Jordan [20]. A previous study has shown high levels of awareness but miscommunication and mistrust leading to low acceptance of donations within the healthcare system. In addition, Lack of organized instruction and truthful communication about death helps reinforce such misconceptions [10]. Addressing These barriers would greatly increase public acceptance of organ donation through focused information campaigns and religious debate.

Emphasizing the critical impact of family and societal expectations in shaping students’ perceptions, the attitude of family value domain obtained the lowest score. That students expressed concerns regarding family rejection is consistent with Jordanian cultural norms where familial consensus often informs medical decisions. Very often, organ donation is considered a decision more appropriately left to older individuals, a mindset that makes younger individuals reluctant.

Social taboos in general regarding dying and bodily integrity might hinder discussions on post-mortem organ donation, leaving the students uncertain if this decision will be acceptable for their family. The management of such uncertainty, on the other hand, may require culturally correct information campaigns where religious opinions regarding organ donation support it, including family discussions. Studies in Jordan confirm that familial and social expectations play an important role in organ donation attitude, and concern about family disapproval and a lack of awareness are major obstacles [10,20]. These may be improved by culturally sensitive awareness initiatives that can increase acceptance and donor registration.

Apart from this, the attitude of distrust of the domain of evaluation reflects latent mistrust in the attitude of the healthcare system regarding the ethical issues of transplantation and organ allocation. Concerns over early removal and its risks to recipients indicate uninformed publics about medical practices that ensure the moral procurement of organs. Past instances of medical negligence or information disparities could enhance distrust and subsequently raise concerns over donation surgery. The need for ethics development along with publicity and the sharing of truthful information related to the selection criteria and methods behind organ allocation will go toward erasing doubts and increasing public faith in the medical fraternity. previous studies in US [23] and Jordan [10,12,21,22] highlight mistrust in the healthcare system, misconceptions about brain death, and ethical issues as significant barriers to organ donation. Indeed, targeted education, transparency in organ allocation, and policy changes could help build public confidence in donation programs.

According to the demographic study, attitudes toward organ donation varied greatly with respect to marital status and age, being more positive among married and older students. Better attitudes among married students could be related to greater exposure to health-related topics in family contexts when the responsibility of making medical decisions becomes clearer. Similarly, the favorable attitudes of the students 26 years of age and above hint at the reality that life experience and maturity improve one’s abilities to understand all the ethical and medical implications involved in organ donation. Younger students, not having had time to experience and directly interact as much with the health problems themselves or ethical questions, are thus more likely to have moderate and negative sentiments. As clarified by many studies, it was confirmed that older and married participants have more positive attitudes toward organ donation due to more life experience and exposure to medical decision-making [25,26]. Yet, cultural moderators might occur that could explain the non-significant correlation between these variables in Iran [27].

The clearest correlation was between attitude scores and willingness to donate; students willing to donate had distinctly more favorable views of organ donation. This finding underlines the closeness of the link between general attitude and personal commitment, hence supporting the need for interventions enhancing willingness through targeted education and exposure to real-life donating situations. Organ donation awareness programs with stories of donors and recipients may help in bridging the gap between conceptual willingness to donate and actual willingness to donate. Research indicates that the propensity to donate is significantly associated with positive attitudes, whereas focused instruction, exposure to actual donations, and previous giving experiences augment donation intentions [20,28,29,30].

The prediction model confirmed that age and willingness to contribute were the most important variables, and taken together, they accounted for 16.0% of variance in opinions. Although readiness to donate came up as the strongest predictor, the relatively low R-squared value reflects that other factor such as religious beliefs, personal experience with organ donation cases, and trust in the health system play very important roles in shaping views. The absence of married status from the final model indicates that its effect is mediated through the other factors and, thus, reinforcing the hypothesis that greater social and personal events define attitudes more than demographic traits alone. Religious beliefs, cultural norms, awareness, and family influences are indeed overwhelming factors in studies of attitudes towards organ donation, often more significant than demographic ones like age and willingness to donate [31,32].

Public policy and health interventions are deeply dependent on these findings. These campaigns must then be designed in a way to target concerns among the young and the unwilling donor, since willingness to donate and age are foremost indicators. The modification of attitude might be mostly dependent on public discussion of the ethical harvesting of organs, social support from relatives, and religious support for organ donation. Moreover, building trust in the healthcare system through more equitable organ allocation policies and ethical safeguards will minimize uncertainty.

This research underlines, above all, the necessity of targeted, culture-specific educational interventions to alter Jordanians’ attitudes towards organ donation. Through structured awareness campaigns, family rejection issues, ethical conduct, and medical trust could be substantially increased levels of acceptance and willingness to donate. More profound psychological and social attitude determinants need to be investigated in subsequent research to develop more comprehensive policies for enhancing wider public participation in organ donation programs.

Targeted policy changes will help to better improve organ donation rates in Jordan. This would include adding organ donation awareness into the core of medical and health-related courses, enabling the understanding and clarification of misconceptions early, while building positive attitudes, this recommendation aligns with multiple studies [33,34,35]. Additionally, this would make it easier for individuals to register as donors through a national registry led by the Ministry of Health, whereas religious concerns and ignorance curtail public participation in Jordan, national donor registries around the world, as seen in Spain and the Saudi Arabia [36], where easy registration procedures facilitate participation, have increased organ donation rates. Clearly targeted policy measures are needed in Jordan, even with the electronic donor register that the Ministry of Health maintains. Thirdly, improved transparency in organ allocation via open regulatory frameworks and publicity mechanisms will eliminate public mistrust and give them confidence in the healthcare system [37]. Similarly, culturally sensitive media campaigns with the help of religious and community leaders are going to lead to education on Islamic perspectives of organ donation, thus facilitating discussions among families [38]. Finally, legal and financial security for the donor families may include covering burial expenses or other benefits [39].

## Conclusion

This study identifies the key factors that significantly influence the attitude of university students towards organ donation in Jordan, including willingness to donate, age, and cultural influences. While many students realize the lifesaving power of organ donation, family approval, religious convictions, and mistrust of organ distribution remain controversial. Targeted legislative actions are thus extremely important to extend organ donation rates. This should encompass integrating organ donation awareness into university curricula, the creation of a national donor registry, culturally sensitive media campaigns in conjunction with religious leaders, and transparency of organ allocation through clearly defined legal frameworks. By implementing these, public trust and organ donation rates could be radically improved, thereby addressing Jordan’s donor shortage.

### Recommendation

Specific education programs need to be incorporated into university curriculums to deal with ethical issues and remove misconceptions, thereby increasing the acceptability of organ donation. Public campaigns featuring testimonials from actual donors and recipients close the divide between theoretical support and actual donation willingness. Well-respected community leaders need to lead religious and cultural discourse removing misconceptions and highlighting religious affirmations of donation. By openness in healthcare and ethical safeguards, public trust in the organ distribution system may be developed. Finally, family-centered initiatives should promote open dialogue about organ donation to balance personal decision-making with social and familial acceptance.

## 5- Limitations

This study has several limitations: cross-sectional design does not allow the establishment of causality between attitudes and influencing factors. The sample consists mainly of university students, which limits the generalization to the broader population. Further, self-reported data are subject to social desirability bias, which may affect the accuracy of responses. Selection bias might have occurred in the way the study was conducted on the social media channels through the elimination of students without access and implying that future studies should be done offline for a representative sample.

## Data Availability

All relevant data are within the manuscript

## Acknowledgment

We acknowledge the equal contributions of the first and second authors in this study and thank all participants for completing the questionnaires.

## Notes

### Competing Interest Statement

The authors have declared that no competing interests exist.

### Clinical Trial

NA

### Funding Statement

The author(s) received no specific funding for this work.

### Author Declarations

The study was approved by the Institutional Review Board at Hashemite University, Zarqa, Jordan.

